# Antibodies to Hepatitis B core antigen prevalence study in Kazakhstan

**DOI:** 10.1101/2022.10.31.22281743

**Authors:** Tatyana Savchuk, Yelena Grinvald, Mohamed Ali, Ramune Sepetiene, Saniya Saussakova, Kuralay Zhangazieva, Dulat Imashpayev, Saniya Abdrakhmanova

## Abstract

**Background:** Kazakhstan is being considered as one of the countries that is medium-endemic for Hepatitis B virus infection. This cross-sectional study aimed to evaluate the prevalence of anti-HBcore and the risk factors impacting positive anti-HBcore markers among donors at Scientific-Production Center of Transfusiology, of the Kazakhstan Republic.

**Study Design and Methods:** The samples taken from blood donors were tested for anti-HBcore, by CLIA on the Architect i2000SR (ABBOTT). In case of positive anti-HBcore, the blood samples were further tested for anti-HBs by CLIA on the Architect i2000SR (ABBOTT). Statistical analysis was conducted by the R software (version 4.1.1, USA, 2021).

**Results and Discussion:** 5709 people aged 18 to 66 years included in the study, the proportion of men and women was 68.17% and 31.83%. The average age of the participants was 35.7±10.57 years.

The prevalence of anti-HBcore among donors was 17.2% (983). Among participants with elevated ALT (170) this marker was determined in 23%, and in donors with normal levels of ALT (5539) - 17%. Participants with positive anti-HBcore scores were on average older (41.8 vs 34.4 years, p<0.001) and Kazakh (88.7% vs 83.0%, p<0.001) by Nationality than study participants with negative results respectively. Anti-HBcore (Antibodies to Hepatitis B core antigen) prevalence in Kazakhstan (17.2%) compared to other countries (8% and more) remains above average. Given the prevalence of HBV and risk factors, it is recommended to include an additional anti-HBcore marker in the mandatory screening of donated blood in the country and improve preventive measures to HBV transmission.

## INTRODUCTION

Hepatitis B (HBV) is a contagious liver disease that results from infection with the Hepatitis B virus. The World Health Organization (WHO) estimated that approximately 2 billion people are carriers of HBV worldwide [1]. Results of epidemiological studies have shown that about 240 million patients have chronic hepatitis B infection (CHB) [2-3].

HBV remains the infection transmissible by direct exposure to infected blood or organic fluids. The existing risk of transmission of HBV is associated with transfusion of blood taken from infected individuals during the window phase and presence of occult HBV [4-7]. High risk of transfusion infection stimulates the realization of the measures for increasing the safety of blood [8].

HBV screening programs often only test for Hepatitis B surface antigen (HBsAg) and antibody to Hepatitis B surface antigen (anti-HBs), not counting those people in whom antibody to hepatitis B core antigen is a positive marker.

Anti-HBcore is produced in infected individuals, both in chronic carrier state and at the end of an acute resolving infection [9]. Also, it is considered an effective marker for occult HBV infection and is an integral part of blood donor screening in many countries.

Alanine aminotransferase (ALT) is a surrogate marker for viral hepatitis. Anti-HBc and ALT tests have been used in Brazil since 1993 [10] and Hungary since 2006 [11]. Results of a 10-year surveillance programme in Germany [12] showed that anti-HBc screening introduced in 2006 has improved blood safety in the country. Study in Netherlands [13] showed similar results. Since 2011 anti-HBc testing has enhanced the safety of the Dutch blood supply but its exact yield remains difficult to determine, due to the complexity of confirming anti-HBc reactivity and occult hepatitis B infection (OBI).

The study by Mosley J. et al. [14] demonstrated that anti-HBc screening would reduce the amount of HBV to be processed by virus inactivation and increase the content of anti-HBs in plasma pools in the USA.

Similarly, Souan L. et al. [15] showed the importance of screening for anti-HBc to improve blood and platelet safety in Jordan. Study carried out by Rodella A. et al. [16] reviewed the importance of the quantitative determination of anti-HBc in provision of additional information and may be useful in the differential diagnosis of acute and chronic HBV infections and in the follow-up of chronically infected patients.

Kazakhstan is ranked as one of the countries with medium endemicity (2-7%) [17]. In 2020 about 23000 HBV cases were registered, the peak incidence was defined in West Kazakhstan, Kyzylorda region and Nur-Sultan city [18]. Approximately 21000 patients have CHB. In contrast, in Kazakhstan anti-HBcore screening for blood donors is not included.

The objective of this study was to determine the prevalence of anti-HBcore among the healthy population and the risk factors impacting positive anti-HBcore markers.

## MATERIALS AND METHODS

### Study design

A cross sectional study was conducted in 2021 in the Scientific-Production Center of Transfusiology of the Ministry of Healthcare in Kazakhstan. Consent was obtained from each prospective blood donor recruited into the study and approval was obtained from Ethics commission (Decision of EC #5 from 20 August 2020).

Participants who met the criteria as blood donors i.e., age between 18 and up, no blood donation in the last 3 months. Those who did not meet the criteria for blood donation stated in the inclusion criteria. The samples taken from blood donors were tested for anti-HBcore, by CLIA on the Architect i2000SR (ABBOTT). In case of positive anti-HBcore, the blood was further tested for anti-HBs by CLIA on the Architect i2000SR (ABBOTT) (Figure 1).

**Fig 1.**
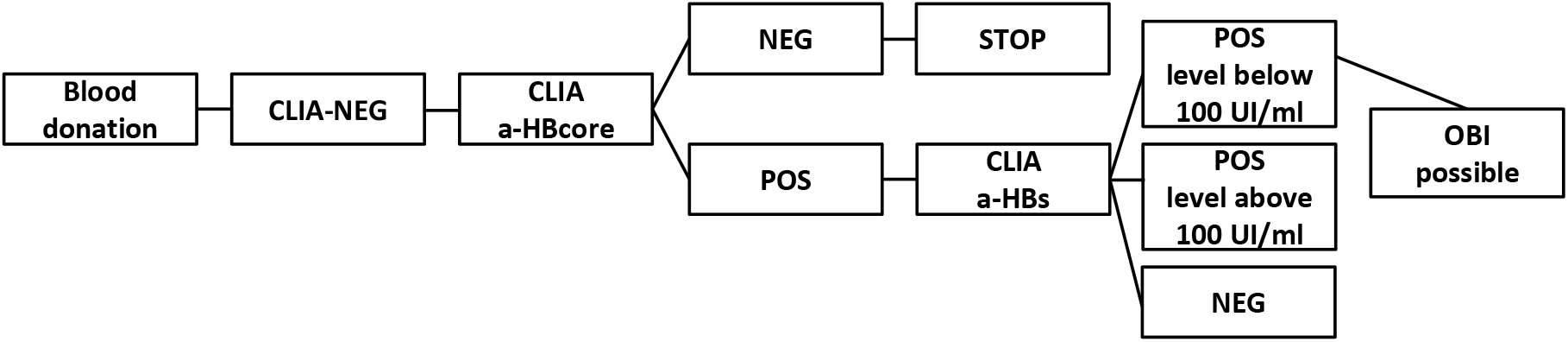
Algorithm for testing donors for the anti-HBcore

ALT were tested by kinetic method on the Biosystems A25. The presence/absence of Human immunodeficiency virus (HIV), Hepatitis B infection DNA (HBV DNA) and Hepatitis C infection were evaluated by testing, through polymerase chain reaction (PCR) techniques in pools of six samples using the Cobas TaqScreen MPX Test v.2.0 multiplex test.

### Statistical analysis

Statistical analysis was conducted by the R software (version 4.1.1, USA, 2021) [19]. Descriptive statistics were reported as proportions (%) for categorical variables and as means±standard deviations or medians with interquartile ranges in parentheses for continuous variables. The normality of the distribution was tested using the Kolmogorov-Smirnov test. Chi-square or Fisher’s exact tests for categorical variables and Student’s T-test or Mann-Whitney U test for continuous independent variables were conducted, as appropriate. Statistically significant results were considered values below p≤0.05.

## RESULTS

A total of 5709 participants was consecutively enrolled in this study. The sample was composed of 68.17% (3139) males and 31.83% (1466) females. The average age of the participants was 35.69±10.57 years ranging from 18 to 66 years. The majority of the participants were of Kazakh origin (84%).

Analysis of anamnestic data showed less than 2% of them had a family member with hepatitis in the last 6 months, and more than 1% of respondents had transfused donated blood or its components in the last 12 months (1.2%),

The prevalence of anti-HBcore among study participants was 17.2% (983). The highest prevalence of anti-HBcore was observed among the young age group 30-39 years old (Fig 2).

**Fig 2.**
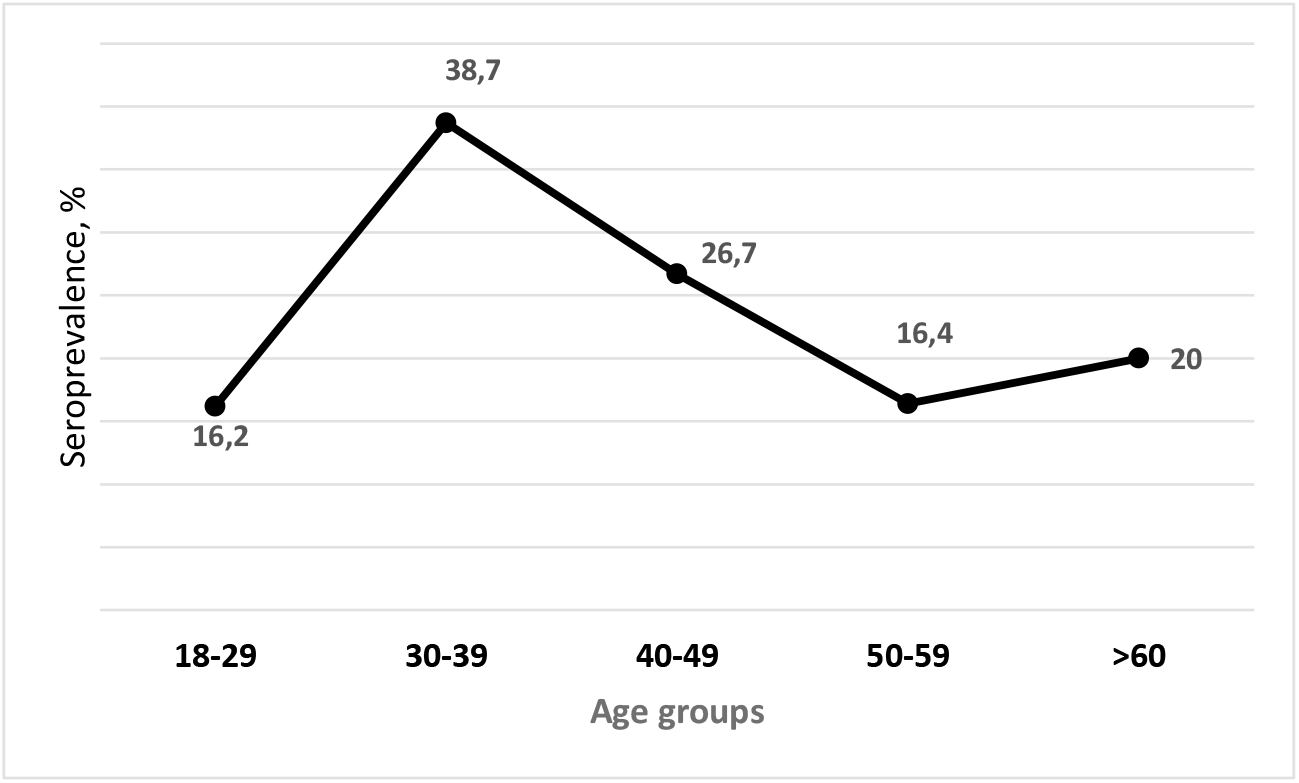
Age-specific distribution of anti-HBcore seroprevalence

Participants with positive anti-HBcore scores were on average older (41.8 vs 34.4 years, p<0.001), Kazakh origin (88.7% vs 83.0%, p<0.001) participants with negative anti-HBcore results respectively (Table 1).

**Table 1.** Data for Descriptive Analysis

| Variables | Total<br>N=5709 | anti-HBcore |  | p-value |
| --- | --- | --- | --- | --- |
|  |  | Negative | Positive |  |
|  |  | n=4726 | n=983 |  |
| Age (mean (SD)) | 35.69 (10.57) | 34.42 (10.33) | 41.75 (9.54) | <0.001 |
| Gender, n (%) |  |  |  | 0.46 |
| Females | 1466 (31.8) | 1220 (32.1) | 246 (30.7) |  |
| Males | 3139 (68.2) | 2583 (67.9) | 556 (69.3) |  |
| Ethnicity, n (%) |  |  |  | <0.001 |
| Kazakh | 3855 (84.0) | 3146 (83.0) | 709 (88.7) |  |
| Russian | 415 (9.0) | 364 (9.6) | 51 (6.4) |  |
| Other ethnicities | 320 (7.0) | 281 (7.4) | 39 (4.9) |  |
| Have family members had hepatitis in the last 6 months? n (%) |  |  |  | 0.41 |
| Yes | 68 (1.6) | 59 (1.7) | 9 (1.2) |  |
| Not | 4096 (98.4) | 3367 (98.3) | 729 (98.8) |  |
| Having transfusion of donated blood or its components in the last 12 months, n (%) |  |  |  | 0.57 |
| Yes | 56 (1.2) | 44 (1.1) | 12 (1.4) |  |
| Not | 4653 (98.8) | 3835 (98.9) | 818 (98.6) |  |
| Having intravenous or intramuscular injections, acupuncture, tattoos or piercings in the last 4 months |  |  |  | 0.71 |
| Yes | 247 (5.2) | 201 (5.2) | 46 (5.6) |  |
| Not | 4458 (94.8) | 3678 (94.8) | 780 (94.4) |  |
| Having surgical interventions (including cosmetic surgery or organ removal), n (%) |  |  |  | 0.18 |
| Yes | 722 (15.3) | 608 (15.7) | 114 (13.7) |  |
| Not | 3989 (84.7) | 3273 (84.3) | 716 (86.3) |  |

Missing answers: for gender - 1104 (19.3%); for ethnicity - 1119 (19.6%), for family history of hepatitis – 1545 (27.1%), for blood transfusions – 1000 (17.5%), for Intravenous or intramuscular injections, acupuncture, tattoos or piercings – 1004 (17.6%), for surgical interventions – 998 (17.5%)

Among participants with elevated ALT (170), positive anti-HBcore marker was determined in 23% (39) participants (Figure 3), and in 17% (944) of donors with normal levels of ALT (5539) (Figure 4).

**Fig 3.**
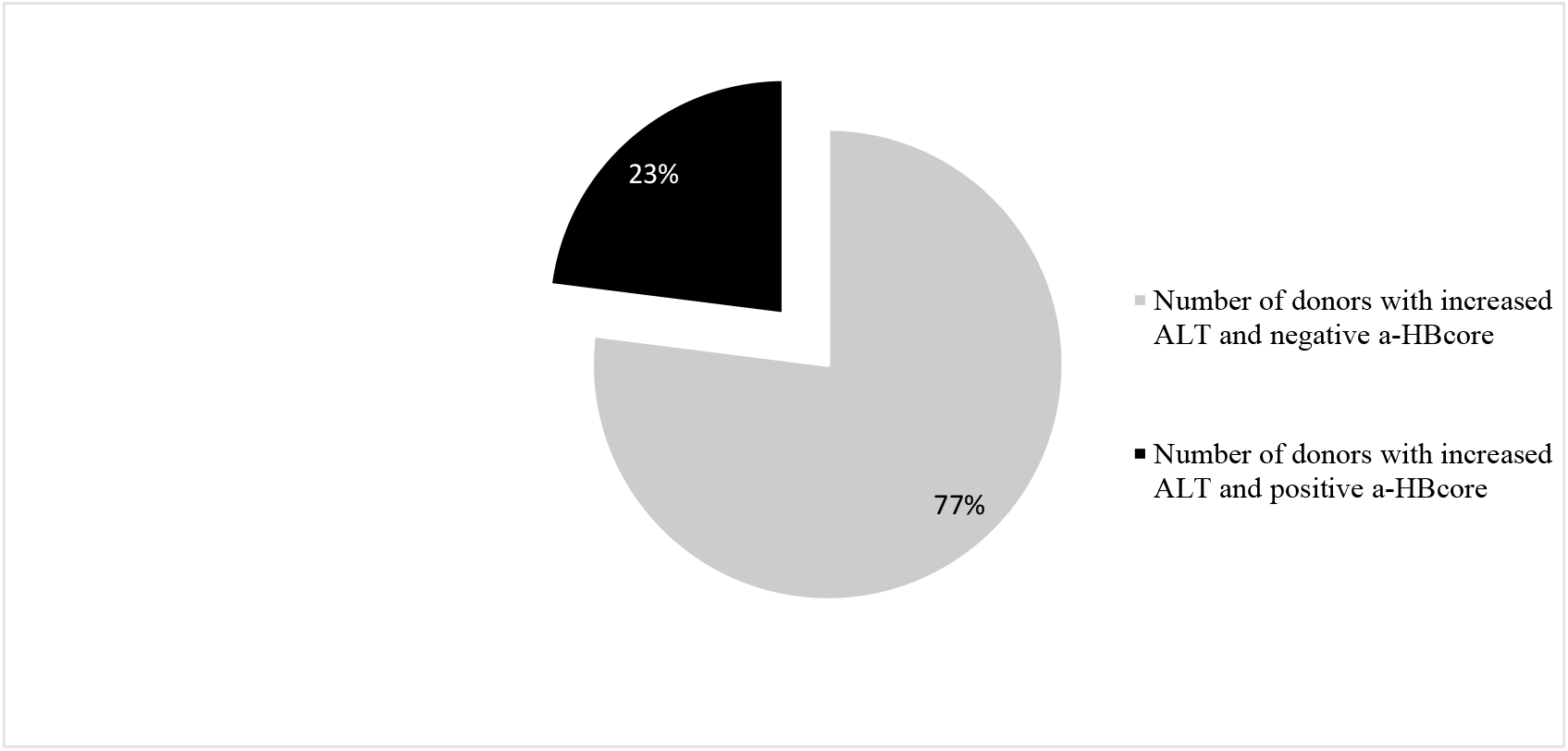
Indicators of a-HBcore detection among donors who had elevated ALT levels

**Fig 4.**
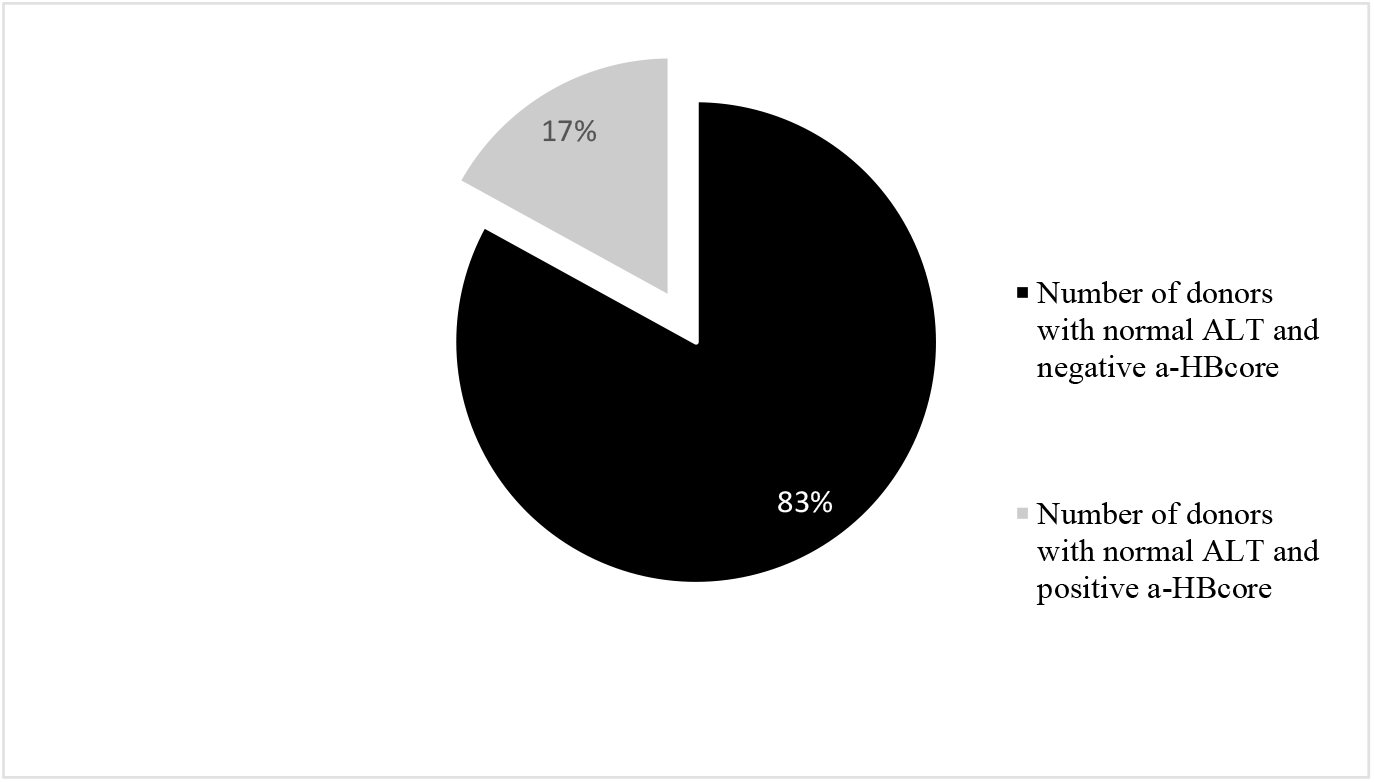
Indicators of a-HBcore detection among donors who had normal

Our interpretation of the results was done according to World Health Organization (WHO) recommendations [20]: positive anti-HBcore marker and anti-HBs marker obtained less than 100 m-IU/ml evaluated as an estimate of discarded donations. Results of testing of anti-HBcore positive samples for the presence of antibodies of HBsAg demonstrated that, 89% (875) have anti-HBs positive markers (Fig.5). The number of donors with a titer of less than 100 m-IU/ml was 298.

**Fig.5.**
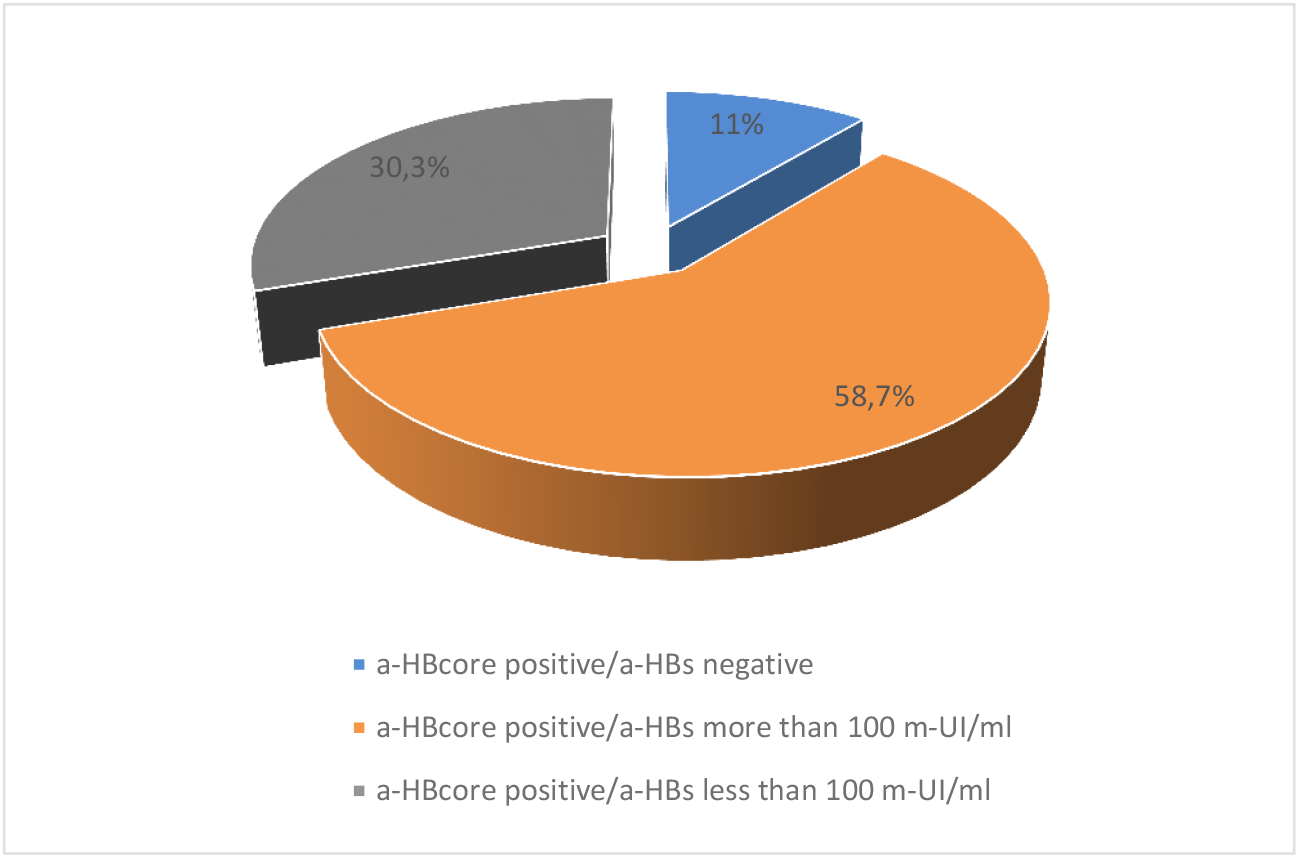
Prevalence of anti-HBcore and anti-HBs markers

## DISCUSSION

We have investigated the seroprevalence of anti-HBcore and anti-HBs among healthy population – blood donors in Kazakhstan. The prevalence of anti-HBcore among the study population was found to be 17,2%. Results of testing of anti-HBcore-positive samples for the presence of antibodies to HBsAg indicated that 89% donors have positive anti-HBs marker.

Houareau C. et al. [12] estimated 0.22% of screened donations were anti-HBcore reactive but HBsAg negative. Study conducted by Villar L. et al. [21] indicated overall prevalence of anti-HBcore and anti-HBs 14.5%, 40.9%, respectively. Anti-HBcore prevalence was positively associated with older age (41 years and above). In our study, the seroprevalence of anti-HBcore was low at the young age group, then peaked among 30-39 years old. We can assume that it might be reasoned due to the introduction of universal HBV vaccination in 1998 [22].

The prevalence of hepatitis B core antigen was 20% in Malaysia [23]. The risk of anti-HBcore seropositivity significantly increased with age and was 53% higher among males than females. Our study also revealed that HBV has more prevalence in males. However, according to the results of a study conducted in Almaty among Russians and Kazakhs, total anti-HBcore was similar in women and men [24]. 9.8% of patients were tested to be positive for anti-HBcore alone in India [25].

Ye X. et al. [26] demonstrated the rate of anti-HBcore positivity increased steadily with age, ranging from the age group younger than 30 years to the age group up to 50 years. A little over 70% of patients carried anti-HBs which is similar to our results. Younger donors with mean age of 25 years were exposed to HBV to a lesser extent compared to those with a mean age of 29 years in study in Pakistani blood donors [27]. Results of study carried out by Muselmani W. [28] suggest including anti-HBcore as an additional screening test for blood donors in Syria to reduce the risk of HBV transmission. In Spain, the rate of anti-HBcore positive cases, HBsAg-negative carriers is approximately 10% of adults between the age of 26 and 65 years [29].

Aguia J. et al. [30] suggest some 33 to 50 percent of cases of hepatitis B that could be transmitted by transfusion of blood from HBsAg-negative donors are prevented by anti-HBcore screening [30]. In Jordan among donors, the prevalence of anti-HBcore was 6.04% [12].

Zhou J. et al. [31] demonstrated that serum anti-HBcore is significantly corrected with hepatic inflammation in CHB patients with normal ALT. In our study, a positive anti-HBcore marker was determined in 23% participants with elevated ALT, and in 17% of donors with normal levels of ALT. Before the NAT era implementation, ALT was widely used in routine practice and called to be a surrogate hepatic infection marker. Despite various updated testing algorithms, ALT value for the detection of hepatitis or any other liver inflammation in its early stage is under debates, otherwise more positive opinions than negative outcomes for clinical prove has no doubts [32-33]. This study has several strengths. The large sample size (5709 participants) allowed for higher statistical power to measure prevalence with greater precision.

Anti-HBcore prevalence in Kazakhstan (17.2%) compared to other countries (Croatia 7%, France 7%, Germany 9%, Iran 16%, Malaysia 20% respectively) remains above average. Given the prevalence of HBV, it is recommended to include an additional anti-HBcore marker in the obligatory screening algorithm of donated blood in the country and improve preventive measures for HBV transmission.

## Data Availability

All original data are available at the Scientific and Production Center of Transfusiology of the Ministry of Healthcare in Kazakhstan.

## Declaration of conflicting interests

The authors declare that there is no conflict of interest in the present study.

## Funding

This research was supported by a grant from Abbott Laboratories, Chicago.

## Author contribution statement

All authors were equally involved.

## Notes

### Competing Interest Statement

The authors have declared no competing interest.

### Funding Statement

This research was supported by a grant from Abbott Laboratories, Chicago, USA

### Author Declarations

Scientific and Production Center of Transfusiology of the Ministry of Healthcare in Kazakhstan, Ethics commission decision of EC#5, 20 August 2020

